# The evolution of knowledge on genes associated with human diseases

**DOI:** 10.1101/2021.06.16.21259049

**Authors:** Thomaz Lüscher-Dias, Rodrigo Juliani Siqueira Dalmolin, Paulo de Paiva Amaral, Tiago Lubiana Alves, Viviane Schuch, Glória Regina Franco, Helder I Nakaya

**Affiliations:** Department of Biochemistry and Immunology, Institute of Biological Sciences, Federal University of Minas Gerais, Belo Horizonte, MG, Brazil; Bioinformatics Multidisciplinary Environment—BioME, IMD, Federal University of Rio Grande do Norte, Natal, RN, Brazil; Department of Biochemistry, CB, Federal University of Rio Grande do Norte, Natal, RN, Brazil; Department of Clinical and Toxicological Analyses, School of Pharmaceutical Sciences, University of São Paulo, São Paulo, Brazil; Hospital Israelita Albert Einstein, São Paulo, Brazil; Scientific Platform Pasteur-University of São Paulo, São Paulo, Brazil

**Keywords:** meta-research, evolution of knowledge, genes, human diseases, network analysis

## Abstract

Thousands of scientific articles describing genes associated with human diseases are published every week. Computational methods such as text mining and machine learning algorithms are now able to automatically detect these associations. In this study, we used a cognitive computing text-mining application to construct a knowledge network comprised of 3,723 genes and 99 diseases. We then tracked the yearly changes on these networks to analyze how our knowledge has evolved in the past 30 years. Our approach helped to unravel the molecular bases of diseases over time, and to detect shared mechanisms between clinically distinct diseases. It also revealed that multi-purpose therapeutic drugs target genes which are commonly associated with several psychiatric, inflammatory, or infectious disorders. By navigating in this knowledge tsunami, we were able to extract relevant biological information and insights about human diseases.

## Introduction

Thousands of scientific articles are published every day, piling up with millions of already published papers (Fortunato et al., 2018). Keeping abreast of scientific significance has become an overwhelming task for researchers in their own fields and in other areas of science. In this scenario, computational methods such as text mining, machine learning, and cognitive computing are helping scientists to summarize published scientific literature. Recently, machine learning approaches have been used to analyze and integrate a variety of biological and medical data (Littmann et al., 2020; Zitnik et al., 2019). These include methods that integrate electronic health records (Rajkomar et al., 2018), capture latent knowledge from the material science literature (Tshitoyan et al., 2019), and discover potential novel drugs to treat psychiatric and neurological disorders using cognitive computing and network medicine analysis of the medical literature (Lüscher Dias et al., 2020).

Particularly, the field of molecular biology has seen a remarkable increase in the number of new studies in recent decades. This has resulted in a large number of genes associated with diseases. As a positive consequence of this efflux of genetic knowledge, diseases that were previously not known to have common etiologies are now being connected through their shared alterations in gene expression and interaction patterns, which has opened many potential new roads for clinical advances (Brooks et al., 2014; Carson et al., 2017; Lees et al., 2011; Postma et al., 2011). One significant example of this trend is the association between psychiatric disorders and immune-related diseases (Gibney and Drexhage, 2013; Marrie et al., 2017; Wang et al., 2015).

Network medicine (Barabási et al., 2011), a contemporary approach to studying relationships between genes and diseases, has also been made possible because of the large amounts of data on genes and diseases available online. Moreover, knowledge networks, that is, complex graphs that connect concepts according to the established knowledge, can be analyzed under the network medicine framework to produce novel insights from medical knowledge (Bai et al., 2016; Lüscher Dias et al., 2020).

In this study, we used IBM Watson for Drug Discovery (WDD; Y. Chen et al., 2016), a cognitive computing text-mining application, to extract known relationships between genes and psychiatric, inflammatory, and infectious diseases from the peer-reviewed literature published between 1990 and 2018. We developed knowledge networks of genes and diseases and monitored the evolution of these relationships yearly. We then quantified and described how genes were connected to each category of disease over this period and how key biological functions unraveled as new genes were added to the network. We also found pairs of diseases from different categories that significantly share genes with each other, indicating underlying clinical proximity between diseases that have not been historically related. Lastly, we explored the genes that were common to all psychiatric, inflammatory, or infectious diseases and investigated which drugs target them. By using a network medicine approach, we were able to extract relevant biological information and new insights of genes, pathways, and therapeutic drugs associated with complex human disorders.

## Results

### Evolution of knowledge on the molecular bases of human diseases

We used WDD, a cognitive computing text-mining application, to identify connections between genes and diseases in millions of peer-reviewed studies (Y. Chen et al., 2016). For each year from 1990 to 2018, we queried WDD to obtain gene sets related to 99 inflammatory, psychiatric, and infectious diseases (Table S1). WDD detects terms of interest, such as genes and diseases, in scientific texts (e.g., PubMed abstracts and full text journal articles) and finds contextual elements connecting them (e.g., prepositions and verbs). These connections can be extracted from many distinct sources of evidence such as gene expression alterations, genome-wide association studies, or protein expression experiments. A confidence score is established for each relationship based on the strength of the detected semantic association and also the number of documents in which the connection is found. However, the type of study from which the association is obtained is not considered for the calculation of the evidence score. Here, we kept only gene-disease relationships with a confidence score equal or higher than 50%, and which were supported by at least 2 studies.

Next, we built yearly disease-disease networks connecting inflammatory, infectious, and psychiatric diseases according to the significance of the genes shared by each pair of diseases (Fig. 1A). These networks were cumulative: the 2018 network (Fig. 1A, rightmost network) displays all connections found in the entire period, while the 2000 network (Fig. 1A, second network from left to right), for instance, contains all connections from 1990 up to that year. The 1990 network (Fig. 1A, leftmost network) depicts the relationships between diseases from the beginning of the literature registries up to 1990.

**Figure 1.**
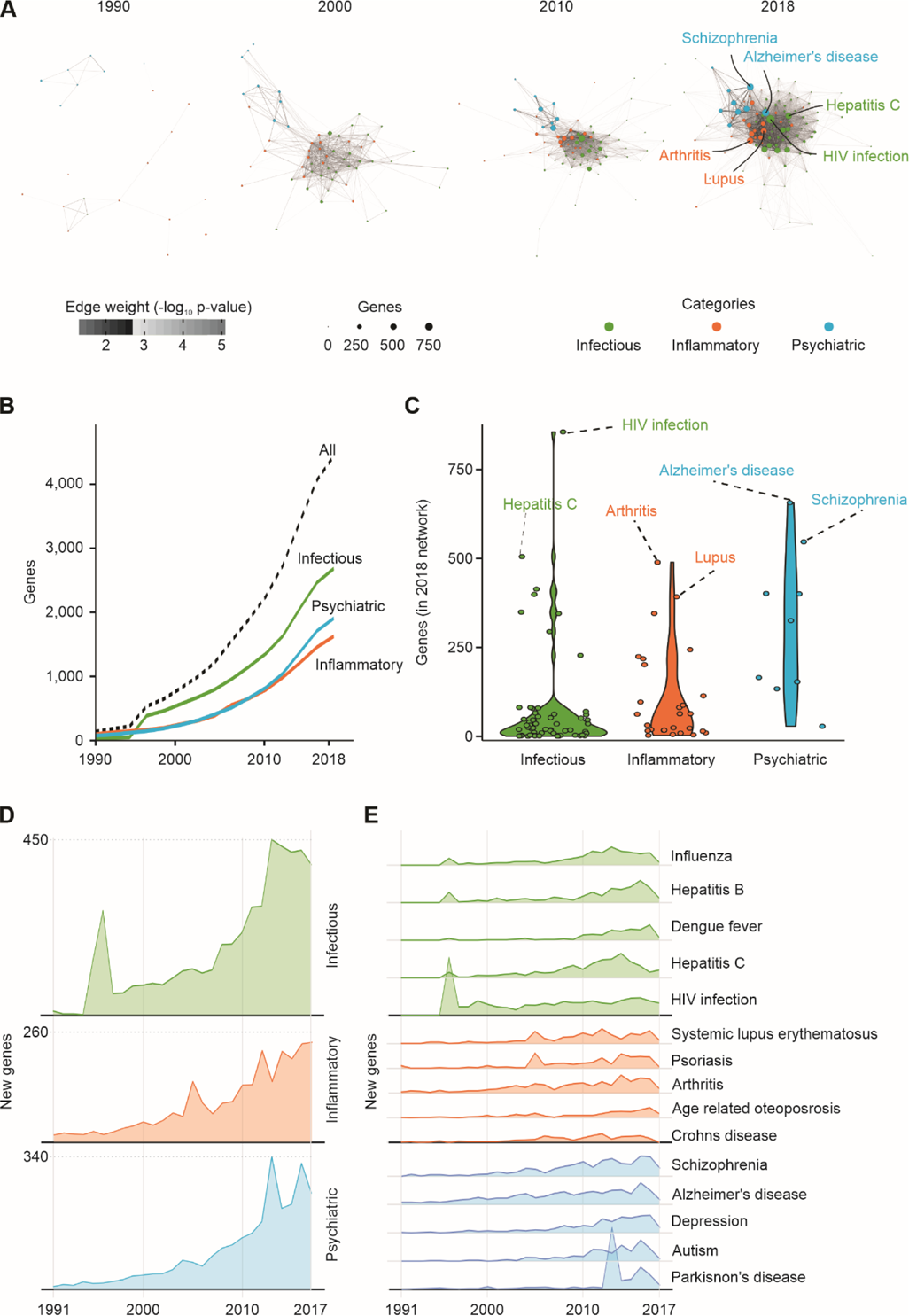
Evolution of knowledge on the molecular bases of human diseases. **A.** Disease-disease knowledge network on infectious, inflammatory, and psychiatric disorders from 1990 to 2018. Nodes represent diseases and are proportional to the number of genes associated with each disease in each year. Edge weights are proportional to the significance of gene-sharing between each pair of diseases. Only edges with a p-value < 0.01 are depicted. **B.** Cumulative number of genes associated with each disease category and with all diseases from 1990 to 2018. **C.** Distribution of the number of genes associated with each disease and category in 2018. **D.** Number of new genes added to the network in each category per year. **E.** Number of new genes added to the network in selected diseases each year. **Color code:** Green – infectious diseases, orange – inflammatory diseases, and blue – psychiatric disorders.

We then assessed how these relationships evolved over the past three decades (1990– 2018) and explored the historical trends of the new genes connected to the network during the period (Fig. 1B–E and Table S2). In 1990, only 95 genes were connected in the network (Fig. 1B), and no association between psychiatric disorders and inflammatory or infectious diseases could be established through shared genes (Fig. 1A). Accordingly, the overall similarity between diseases (between or within categories) was low in 1990 (Fig. S1). From 1990 to 2010, with the constant increase in the number of genes associated with diseases in all categories, a preliminary approximation between inflammatory and infectious diseases was observed (Fig. 1A, second panel, and Fig. S1A). During the next 9 years (2010 to 2018), the new genes added to the network (Fig. 1B) resulted in a strengthening of the connections between infectious and inflammatory diseases, and a fast approximation between psychiatric disorders and the other two categories (Fig. 1A, fourth panel, and Fig. S1A). Meanwhile, the proximity of diseases within the same categories also increased (Fig. S1B). Inflammatory diseases occupy a central position in the 2018 network (Fig. 1A, fourth panel), which reflects their high between- and within-category similarities sustained throughout the 30-year period (Fig. S1). Psychiatric and infectious diseases presented the lowest similarity between each other (Figs. 1A and S1).

In 2018, a total of 3,723 genes were present in the network (Fig. 1B). The number of genes associated with each disease in the three different categories in 2018 also varied (Fig. 1C). The infectious diseases with the highest number of connected genes in 2018 were hepatitis B (414 genes), hepatitis C (506 genes), and HIV infection (856 genes; Fig. 1C). However, 55 of 63 infectious diseases were connected to less than 100 genes in 2018 (Fig. 1C). The most connected inflammatory diseases were psoriasis (346 genes), systemic lupus erythematosus (393 genes), and arthritis (490 genes; Fig. 1C). In the category of psychiatric disorders, Alzheimer’s disease was the most connected (657 genes), followed by schizophrenia (547 genes) and depression (402 genes; Fig. 1B). The imbalance in the distribution of genes connected to infectious diseases likely reflects a bias in the research interest toward the discovery of genes related to diseases already connected to more genes. In fact, the 2018 network showed that the number of scientific papers that mentioned poorly connected diseases (less than 100 genes) is significantly lower than the number of papers published on highly connected diseases (more than 100 genes) (Fig. S1C).

Distinct historical trends of discovery were seen for each disease category (Fig. 1D and Table S3). Prominent peaks of gene-association discovery occurred in 1996 for infectious diseases, in 2005 for inflammatory diseases, and in 2013 for psychiatric disorders (Fig. 1C). From 2010 to 2017, the rate of gene discovery in all three categories increased (Fig. 1C). The significant increase in the number of genes associated with infectious diseases observed in 1996 was mostly driven by 154 new genes associated with HIV infection (Fig. 1D), which corresponded to 50% of the new genes added to the network in that year (Table S3). The triple therapy for HIV using nucleoside reverse-transcriptase inhibitors and protease inhibitors was established in 1996 (Hammer et al., 1996), which likely influenced this outburst of genetic discovery. The 2005 increase in the number of genes associated with inflammatory diseases was mostly related to the new genes connected to psoriasis (41 genes) and systemic lupus erythematosus (33 genes; Fig. 1D), which together corresponded to 20% of the new genes associated with all of the diseases in 2005 (Table S3). The Th_17_ cell lineage was discovered in 2005 (Langrish et al., 2005), a cell type that has since been strongly associated with autoimmune and infectious diseases (Zambrano-Zaragoza et al., 2014). In 2013, a large number of new genes were associated with Parkinson’s disease (165 genes Fig. 1D), which corresponded to 17% of the new genes in the network in that year (Table S3). We could not detect any specific scientific landmark in 2013 that could explain this peak. Nevertheless, important genes related to the innate immune response to pathogens and inflammation are among the new genes associated with Parkinson’s disease in 2013, such as interleukin 1 beta (IL1B) and the p105 subunit of the nuclear factor kappa B (NFKB1).

### Evolution of disease relationships between categories

Next, we investigated the evolution of the similarity between diseases from different categories according to their shared genes (see Methods section). For the top 9 most connected diseases of each category in 2018 (i.e., diseases connected to more genes), we detected the diseases from the other two categories with the most significant gene sharing between them and analyzed how these relationships evolved from 1990 to 2018 (Figs. 2, S2, S3, and S4). Alzheimer’s disease was the psychiatric disorder with the highest similarity to inflammatory diseases in 2018, including arthritis and systemic lupus erythematosus (Fig. 2A). The relationships between Alzheimer’s disease and these disorders grew steadily in significance from 1990 to 2018 (Fig. S2A), which captures the now well-established relevance of inflammatory processes in the pathophysiology of Alzheimer’s disease (Newcombe et al., 2018). Surprisingly, fibromyalgia was similar to several psychiatric diseases: depression, anxiety, bipolar disorder, schizophrenia, and Huntington’s disease (Figs. 2A and S2). The total number of genes associated with fibromyalgia in 2018 was low (25 genes), but 72% of these (17 genes) are also associated with depression. These are genes related to nervous system development, such as brain derived neurotrophic factor (BDNF), nerve growth factor (NGF), and neuropeptide Y (NPY), and inflammatory response, including interleukin 6 (IL6), C-X-C motif chemokine ligand 8 (CXCL8), and tumor necrosis factor (TNF). In fact, fibromyalgia patients often present psychiatric comorbidities such as depression and anxiety (Galvez-Sánchez et al., 2020).

**Figure 2.**
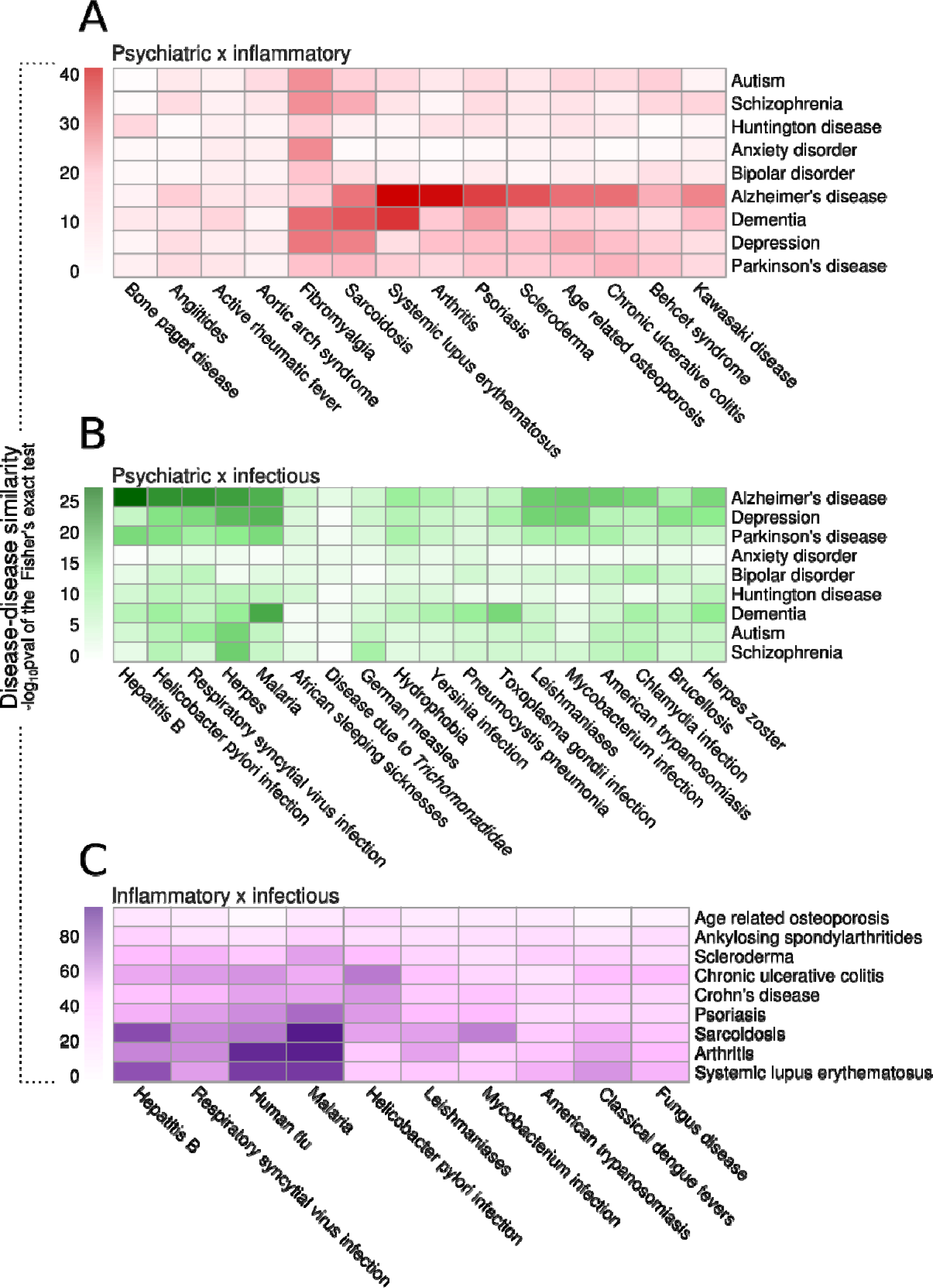
Evolution of disease relationships between categories. **A.** Disease-disease similarity between diseases of different categories in the 2018 network according to their shared genes. The similarity score was defined as the –log_10_pval of the Fisher’s exact test result of the gene overlap between each disease pair. Each heatmap represents the similarity score between diseases of two different categories: **A.** psychiatric versus inflammatory diseases. **B.** psychiatric versus infectious diseases. **C.** inflammatory versus infectious diseases.

**Figure 3.**
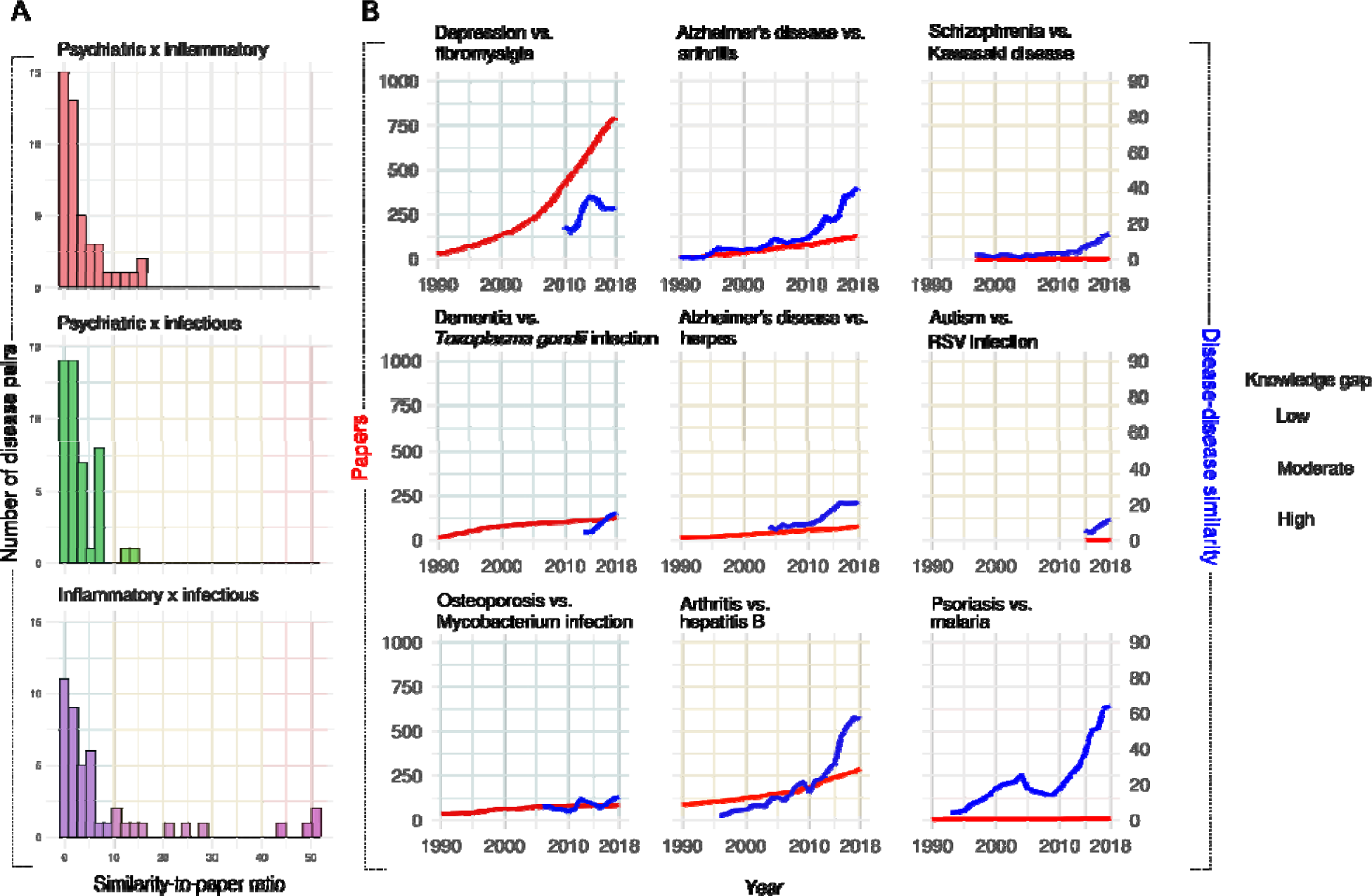
Evolution of the knowledge gap between diseases of different categories. A. Number of disease pairs according to the similarity-to-paper ratio index. This index was obtained as a ratio of the similarity score to the total number of papers published for each disease pair in 2018. Low similarity-to-paper ratio (<10) is colored in blue; intermediate similarity-to-paper ratio (10 < ratio < 40) is colored in yellow; and high similarity-to-paper ratio (>40) is colored in pink. **B.** Selected cases of disease pairs with low, intermediate, or high similarity-to-paper ratios depicting the evolution in the number of papers on each pair and the evolution of the similarity between them.

Among infectious diseases, herpes was the most similar disease to autism, schizophrenia, and Huntington’s disease and was also among the top 5 most similar infectious diseases to depression, Parkinson’s disease, and Alzheimer’s disease (Figs. 2B and S3). Herpes infection might be associated with the development of Alzheimer’s disease (Harris and Harris, 2015); the typical amyloid-β deposition that occurs in the brain of Alzheimer’s disease patients could be an innate immunity mechanism to fight herpes virus infections (Eimer et al., 2018). Our results indicate that there has been latent evidence of that association since the early 2000s in the scientific literature (Fig. S3A). In the 2005 network, Alzheimer’s disease and herpes virus infection shared 14 genes, which represented 58% of the known genes associated with herpes infection at that time.

Autoimmune inflammatory diseases, such as systemic lupus erythematosus, arthritis, and psoriasis, also showed strong gene sharing with viral infections such as hepatitis B and C, respiratory syncytial virus (RSV) infection, influenza, and HIV (Figs. 2C and S4). The association between viral infections and autoimmune diseases is well documented (Getts et al., 2013). For instance, the SARS-CoV-2 virus can trigger Guillain–Barré syndrome, a neurological autoimmune disease, in COVID-19 patients (Dalakas, 2020). Dengue patients also present a higher risk of developing autoimmune diseases, such as systemic lupus erythematosus and vasculitis (Li et al., 2018), an association that was also captured in our analysis of the scientific literature since the late 1990s (Fig. S4I).

We then examined the number of publications retrieved from PubMed using the topmost similar pairs of diseases from distinct categories as queries (see Methods section; Fig. 3). The goal was to find out whether the gene-sharing similarities between diseases from different categories detected in our networks could also be captured from direct co-occurrence in the general peer-reviewed literature over the 30-year period. For each disease pair, we obtained a ratio between the similarity score of the diseases (i.e., the significance of the gene sharing between them) and the total number of studies retrieved from PubMed that mention both diseases of the pairs together (Table S4). This similarity-to-paper ratio was used to detect potentially understudied pairs of diseases that significantly share genes. Low similarity-to-paper ratio values (Figs. 3A and 3B, light green, and Table S4) represent similar diseases with many papers already published about them or dissimilar disease pairs. An example of such a pair is fibromyalgia and depression. These diseases have significant gene sharing and also hundreds of scientific papers that explore their relationship in the literature (Fig. 3B). Conversely, the genetic association between osteoporosis and mycobacterial infection is low and so is the number of papers that investigate these diseases together (Fig. 3B). These cases were considered as examples of a low knowledge gap between the genetic similarity obtained from our network analysis and the established literature coverage of the disease pairs.

Cases with an intermediate similarity-to-paper ratio (Figs. 3A and 3B, yellow, and Table S4) were considered as cases of moderate knowledge gap (Fig. 3A), which was the case for arthritis and hepatitis B (Fig. 3B). As previously mentioned, several recent studies have explored the association between viral infections and autoimmune diseases (Dalakas, 2020; Getts et al., 2013; Li et al., 2018). In 2018, there were over 250 published papers in which arthritis and hepatitis B were mentioned together (Fig. 3B). Virally mediated arthritis represents ∼1% of all arthritis cases, including cases related to hepatitis B infection (Marks and Marks, 2016). Scientists have detected the hepatitis B virus in the synovial fluid of rheumatological patients, which could contribute to the pathogenesis of arthritis (Chen et al., 2018). Although these diseases are known to be clinically associated at least since the 1970s (Mirise and Kitridou, 1979), our results show that the knowledge on the gene sharing between them increased rapidly after 2015, which was not followed at the same rate by the number of papers published on the two diseases together. This represents a potential gap to be explored by novel research on the genetic bases of the relationship between arthritis and hepatitis B.

Lastly, we considered the disease pairs with strong gene sharing and few studies supporting a direct association as cases of a high knowledge gap (Figs. 3A and 3B, pink, and Table S4). We suggest that these cases might represent potentially underexplored fields of research that deserve further investigation. Surprisingly, the number of papers published until 2018 that mentioned psoriasis and malaria together was neglectable (Fig. 3B). These diseases share 31 genes, one-third of the genes associated with psoriasis, and over 10% of the genes associated with malaria in the 2018 network. Hydroxychloroquine, a drug used to treat malaria (Ben-Zvi et al., 2012) and rheumatic diseases, such as arthritis and lupus (Ben-Zvi et al., 2012), can trigger psoriatic lesions (Balak and Hajdarbegovic, 2017). Among a few papers in which malaria and psoriasis are mentioned together, there is a report from 2014 that describes cases of hydroxychloroquine-induced psoriasis in patients undergoing malaria treatment (Gravani et al., 2014). The authors of this study suggest that there should be guidelines for the management of psoriasis patients who are also at risk of malaria (Gravani et al., 2014). Our findings corroborate the need for future studies to investigate the association between these diseases.

### Evolution of biological pathways

We performed a gene overrepresentation analysis (ORA) against Reactome pathways with the genes associated with the top 9 most connected diseases in each year from 1990 to 2018 (Figs. 4-6 and Table S5). We detected 433 Reactome pathways that presented significant enrichment (p.adjust < 0.01) among the genes of at least one disease (Table S5). Functional enrichment analysis, such as ORA, often yields too many significant pathways, making these results difficult to interpret at the individual pathway level. For this reason, we used a network approach to reduce the complexity of the obtained set of enriched pathways (see Methods section). Briefly, we built a pathway network (Fig. 4) with the significant Reactome pathways obtained from the ORA. We connected these pathways to each other according to the gene sharing between them, similar to what was done in Fig. 1A. We then identified 11 clusters of closely connected pathways in the network and annotated these clusters according to the main biological functions of the pathways within them (Fig. 4 and Table S5). One of the detected clusters grouped several pathways associated with interferon-stimulated genes, interleukins, and antigen presentation (Fig. 4 and Table S5). The pathways in this cluster were significantly enriched among the genes of diseases in all categories, including malaria, HIV infection, arthritis, lupus, depression, and Alzheimer’s disease (Fig. 5). The pathways related to interleukin signaling (e.g., “interleukin 10 signaling”), for instance, were among the top enriched pathways associated with depression genes in the 2018 network (Fig. 5 and Table S5). Another cluster of pathways that showed consistent enrichment across all disease categories was NF B-κ mediated inflammation induced by toll-like receptors (TLRs), T-cell receptors (TCRs), and B-cell receptors (BCRs; Fig. 4). These results illustrate the most recurring theme detected in our study: psychiatric, inflammatory, and infectious diseases share common immunological mechanisms that are mostly related to innate immunity and inflammation.

**Figure 4.**
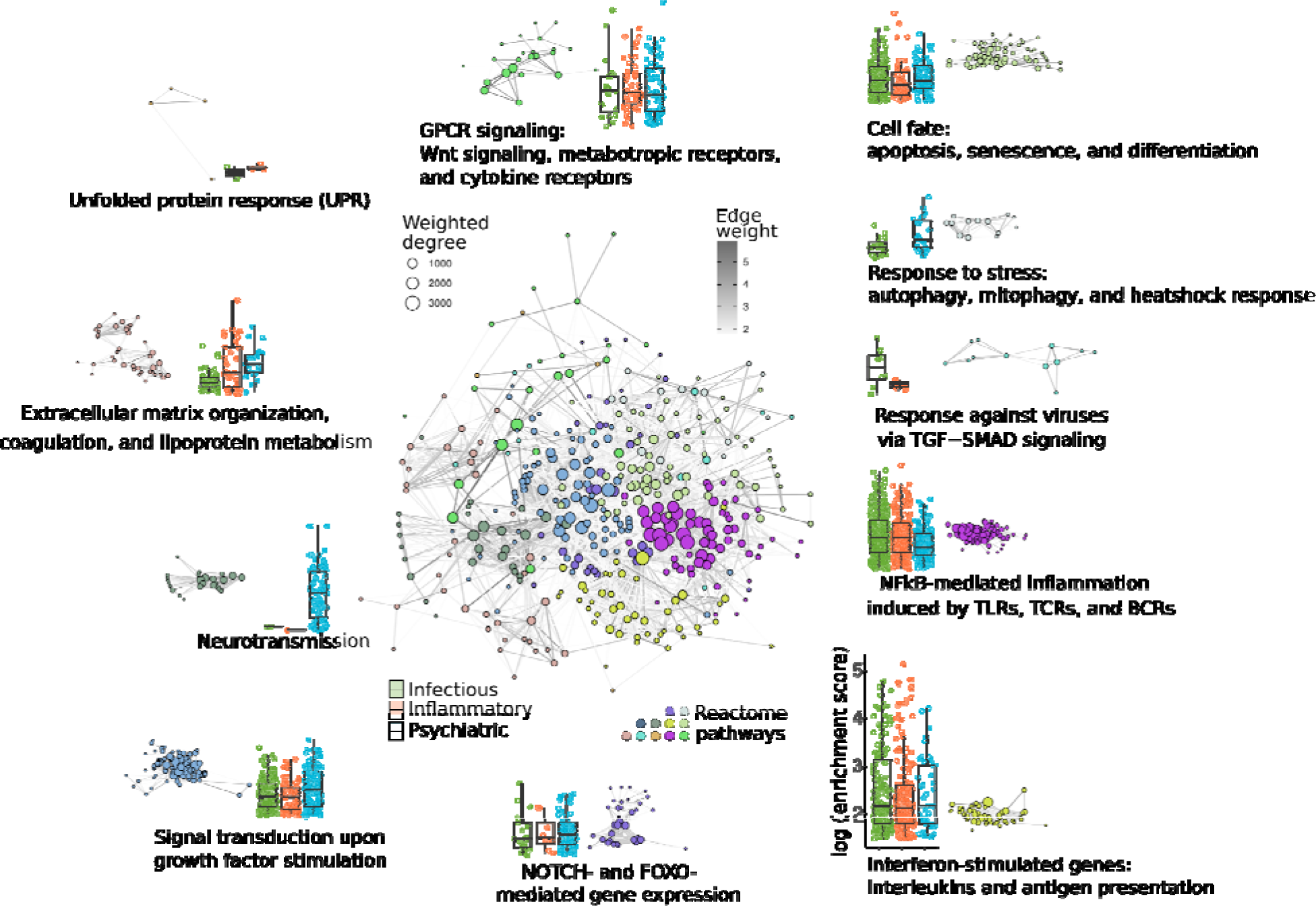
Reactome term network built from the ORA results of the genes associated with human diseases in 2018. Significant Reactome ORA terms (p.adjust < 0.01) obtained from the genes of the top 9 diseases in the 2018 network were connected to each other according to the significance of the gene sharing between them (edge weight). Only terms with a gene sharing with a p.adjust < 0.01 were connected. We detected 11 clusters (node colors) of closely related terms using the Louvain clustering algorithm in the R package *igraph* (Csardi and Nepusz, 2006) and compared the enrichment score distribution of the terms in these clusters in each disease category (box plots). Box plots are colored according to the disease categories: green – infectious diseases, orange – inflammatory diseases, and light blue – psychiatric disorders. Dots in the box plots represent individual enriched Reactome pathways that belong to each network cluster.

**Figure 5.**
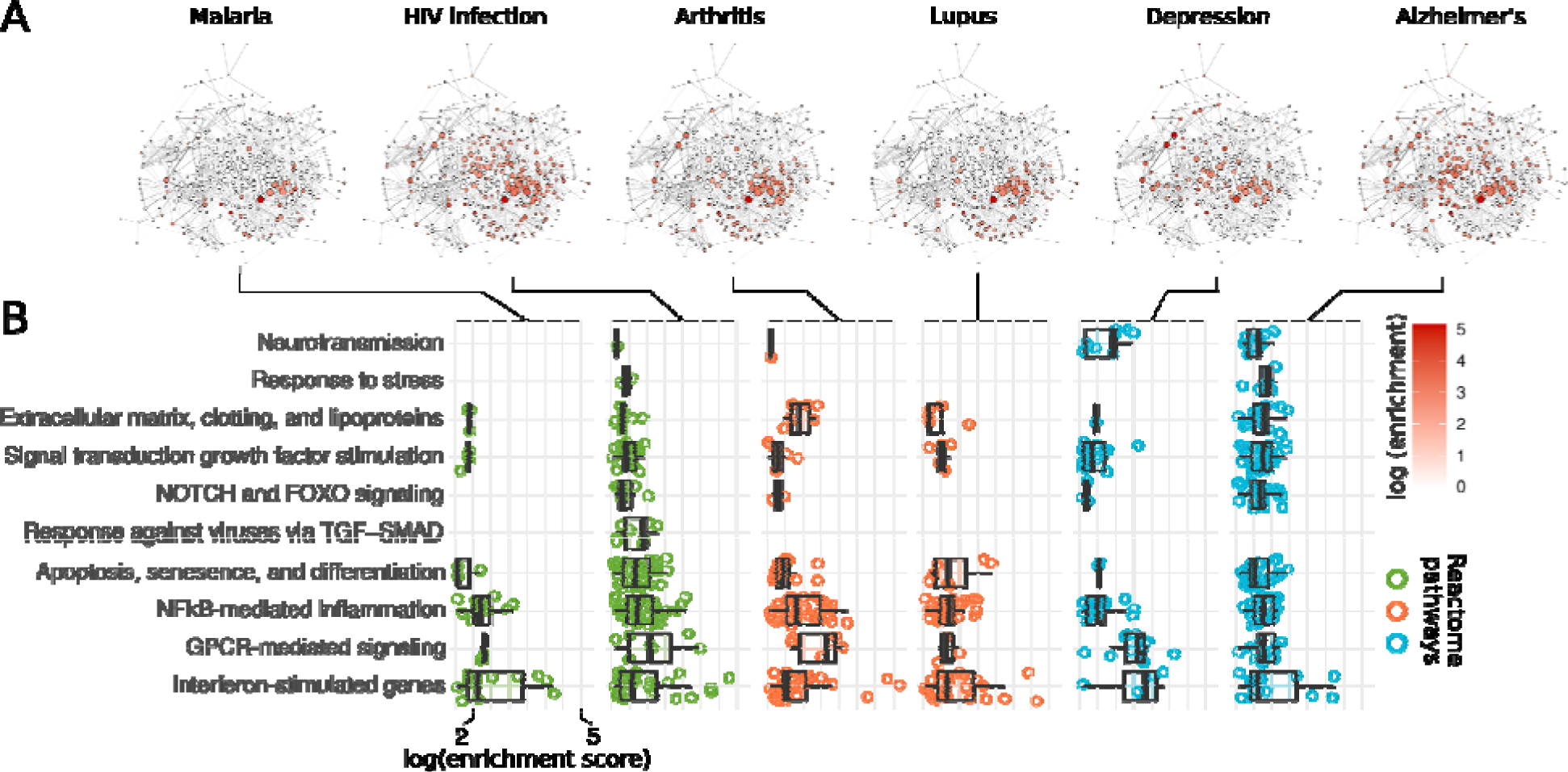
Key biological pathways are enriched among the genes associated with human diseases in 2018. **A.** ORA networks depicting the enrichment score of Reactome pathways in selected infectious, inflammatory and psychiatric disorders. The networks in A have the same topology of the network in Figure 04. The nodes are colored according to the logarithm of enrichment score (-log_10_pval) of the terms represented by each node. **B.** ORA enrichment score distribution of the terms in the clusters and diseases from panel A. Box plots are colored according to the category of each disease: green – infectious, orange – inflammatory, and light blue – psychiatric. Dots in the box plots represent individual Reactome pathways that belong to the clusters listed in the y axis and that were enriched in each disease.

**Figure 6.**
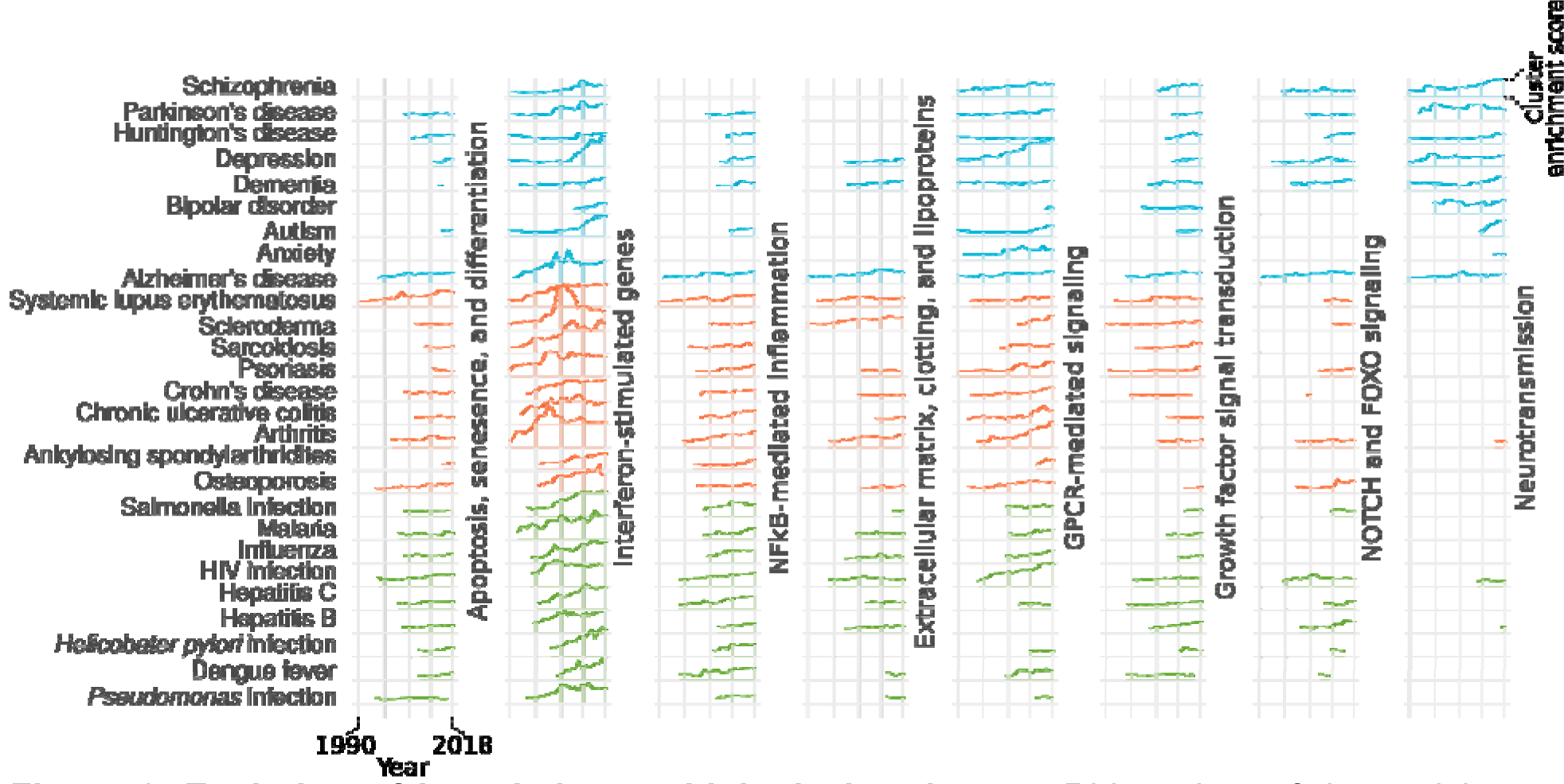
Evolution of knowledge on biological pathways. Ridge plots of the enrichment score of selected clusters from the network in Figure 04 for the top 9 diseases in each category from 1990 to 2018. The height of the ridges are proportional to the mean enrichment score (mean log_10_pval) of the Reactome pathways in each cluster listed in the y axis.

Conversely, we found a cluster of closely connected pathways related to neurotransmission that were enriched mostly among the genes of psychiatric disorders (Fig. 4 and Table S5). However, three inflammatory and infectious diseases (hepatitis B, arthritis, and HIV infection) presented enrichment for pathways in this cluster (Fig. 5 and Fig. S5). The genes related to these diseases presented enrichment for the pathway “transcriptional regulation MECP2”, a member of the neurotransmission cluster. Methyl CpG binding protein 2 (MECP2) is located in the X chromosome, and mutations in this gene are the primary cause of Rett syndrome (Liyanage and Rastegar, 2014). There is no evidence in the scientific literature that there is a link between HIV infection or hepatitis B and Rett syndrome, but recent studies indicate a link between this neurodevelopmental disorder and autoimmune diseases, including arthritis (De Felice et al., 2016). Moreover, AIDS patients can develop neurological manifestations similar to those observed in Rett patients, such as cognitive dysfunction and movement disorders (Brew and Garber, 2018). Our results suggest that the similarity between Rett syndrome and autoimmune diseases might also occur for infectious diseases of viral etiology.

We also detected other clusters of pathways with similar enrichment results between diseases of different categories (Fig. 4). The genes related to arthritis and those related to Alzheimer’s disease presented enrichment for pathways related to the extracellular matrix organization, coagulation, and lipoprotein metabolism (Fig. 5). In arthritis, fibroblast-like synoviocytes become hyper-inflammatory and disrupt the extracellular matrix integrity, which leads to the degradation of synovial joint collagen (Nygaard and Firestein, 2020). In Alzheimer’s disease, some extracellular matrix macromolecules seem to promote the production and stabilization of amyloid β, while others act to protect neurons from amyloidosis (Sethi and Zaia, 2017). The pathways in the signal transduction on growth factor stimulation and GPCR-mediated signaling clusters were also enriched among the genes of diseases in all categories (Figs. 4, 5, and S5). This result was expected because the genes involved in signal transduction and intracellular signaling are usually shared between cellular pathways and are involved in virtually all biological functions relevant to diseases (Figs. 5 and S5).

After determining the major biological functions related to the genes connected to infectious, inflammatory, and psychiatric diseases in the 2018 network, we investigated how this knowledge evolved from 1990 to 2018 (Fig. 6). The pathways related to interferon-stimulated genes, interleukins, and antigen presentation became enriched for the genes associated with inflammatory and infectious diseases already since the early 1990s (Fig. 6). Surprisingly, this enrichment appeared earlier for inflammatory diseases, despite the highly relevant role of interferon-stimulated genes and antigen presentation in infectious diseases. Conversely, there was a significant increase in the enrichment of these pathways for the genes related to depression, autism, and schizophrenia since 2010 (Fig. 6). Recently, the specific roles of the immune system in psychiatric diseases begun to be revealed (Chen et al., 2016; de Baumont et al., 2015; Dong et al., 2018; Madore et al., 2016; Yuan et al., 2019). Particularly, neuroglial cells have gained importance as key neuroimmune players in the development of autism (microglia and oligodendrocytes; Scuderi and Verkhratsky, 2020), Alzheimer’s disease (microglia; Clayton et al., 2017), and schizophrenia (astrocytes; Gandal et al., 2018). The association of pathways related to apoptosis, senescence, and cell differentiation with psychiatric disorders has also occurred recently, except with Alzheimer’s disease, which began early in the period (Fig. 6).

Alzheimer’s, Parkinson’s, and Huntington’s diseases are neurodegenerative conditions in which chronic neuronal death happens in distinct parts of the brain (Dugger and Dickson, 2017). We also found an increasing association in recent years of genes related to autism and depression to cell fate pathways (Fig. 6), showing that these disorders might also have a neurodegenerative component. In fact, apoptosis and cell death in response to stress and inflammation are relevant factors in the pathogenesis of autism (D. Dong et al., 2018) and depression (Leonard, 2018).

### Evolution of drug target hub genes

Lastly, we examined how drugs that are used to treat inflammatory, infectious, and psychiatric diseases target the genes that are shared between the three categories. We found that 345 genes were common to all disease categories (Fig. 7A). Ninety-nine genes were shared only between inflammatory and psychiatric diseases; 259 were common only between psychiatric and infectious diseases; and a total of 409 genes were related exclusively to inflammatory and infectious diseases (Fig. 7A). The remaining genes were unique to inflammatory (493 genes), psychiatric (869 genes), and infectious diseases (1,209 genes; Fig. 7A).

**Figure 7.**
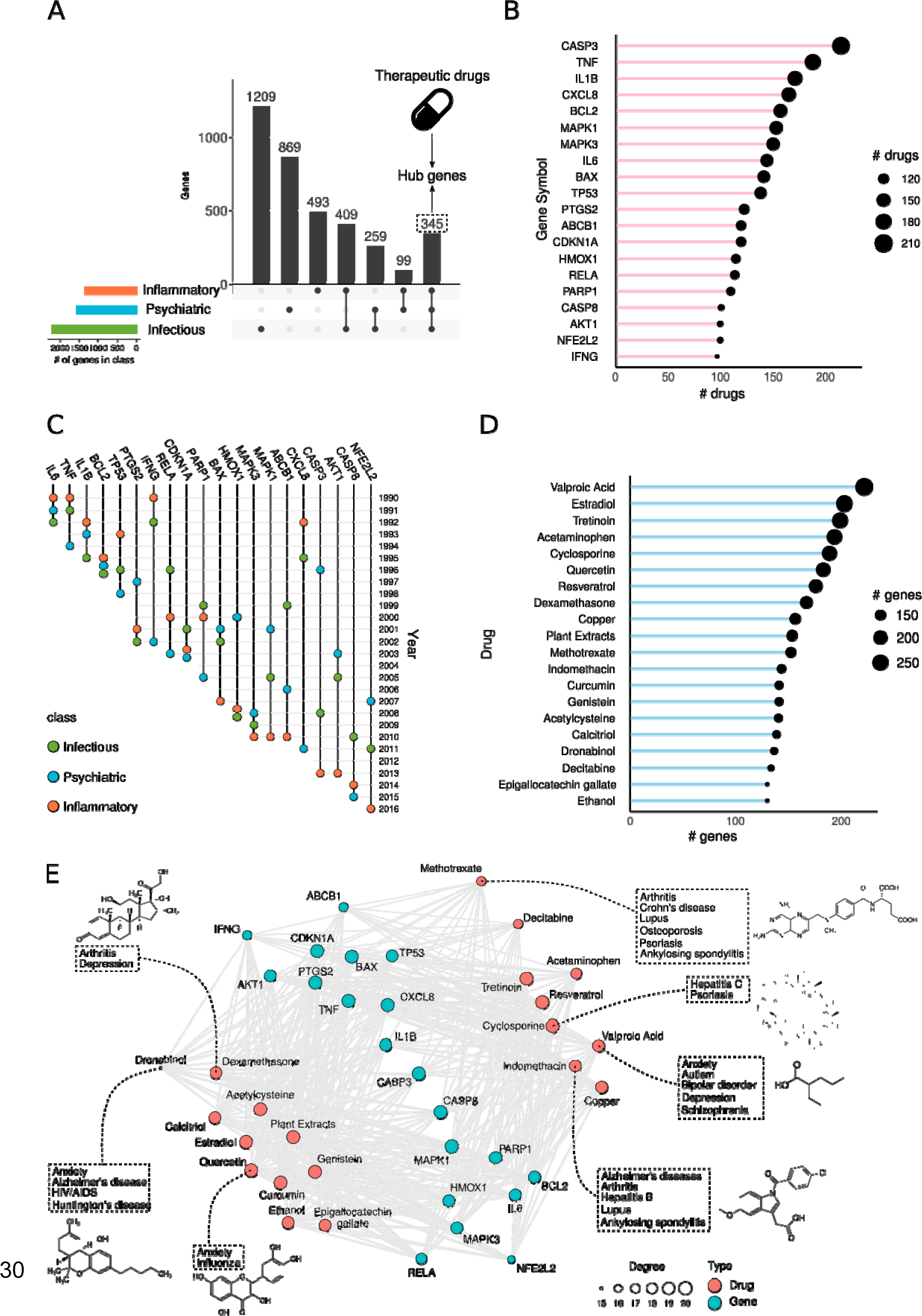
Evolution of drug target hub genes. **A.** UpSet plot showing the common genes between all categories (hub genes), between two categories exclusively and genes that are unique to each category. **B.** Number of therapeutic drugs of inflammatory, infectious, and psychiatric diseases that target the top 20 target hub genes according to the comparative toxicogenomics database (CTD). **C.** Timeline of the association of the top 20 target hub genes to the gene-disease network. The year in which each gene was associated with the first disease of each category is depicted by the circles with distinct colors for each category. **D.** Number of hub genes targeted by the top 20 drugs that target more hubs according to CTD. **E.** Drug-gene network depicting the top 20 drugs and that target hub genes. We selected a few drugs and illustrated their molecular structure and diseases for which they are listed as therapeutic according to CTD.

We used the comparative toxicogenomics database (CTD; Davis et al., 2021) to find drugs that have a therapeutic relationship with the top 9 diseases and the list of genes that these drugs affect (see Methods section). From these lists, we highlight the top 20 most common target genes of the therapeutic drugs listed by CTD (Fig. 7B). Among these genes, IL6, TNF, and interferon gamma (IFNG) were already connected to inflammatory diseases in the 1990 network and were gradually related to diseases in the other two categories until 2002 (Fig. 7C). Interleukin 1 beta (IL1B), B-cell lymphoma 2 (BCL2), tumor protein P53 (TP53), and CXCL8 also appeared in our networks in the early 1990s and were first connected to inflammatory diseases (Fig. 7C). Eight drug target genes were first connected to psychiatric disorders (Fig. 7C): caspase 3 (CASP3; 1996), prostaglandin-endoperoxide synthase 2 (PTGS2; 1997), heme oxygenase 1 (HMOX1; 2000), BCL-2-associated X (BAX) and mitogen-activated protein kinase 1 (MAPK1; 2001), RAC-alpha serine/threonine-protein kinase (AKT1; 2003), nuclear factor erythroid 2-related factor 2 (NFE2L2; 2007), and mitogen-activated protein kinase 1 (MAPK3; 2008). The other 5 genes were first connected to infectious diseases (Fig. 7C): NFkB P65 Subunit (RELA; 1996), poly(ADP-Ribose) polymerase 1 (PARP1) and ATP binding cassette subfamily B member 1 (ABCB1; 1999), cyclin dependent kinase inhibitor 1A (CDKN1A; 2001), and caspase 8 (CASP8 ; 2010). All top 20 drug target genes were first connected to one of the categories until 2010, with the majority of new connections happening in the 1990s (Fig. 7C). These are very well-known genes involved in inflammation (e.g., IL6 and IL1B), innate immunity (e.g., IFNG), apoptosis (e.g., CASP3 and CASP8), cell cycle (e.g., TP53), and other key biological functions that are altered in several diseases.

Next, we found the top 20 therapeutic drugs that affect the most hub genes of inflammatory, psychiatric, and infectious diseases (Fig. 7D). Valproic acid, a class I histone deacetylase (HDAC) inhibitor (Göttlicher et al., 2001), was the drug that affected the most hub genes, 259 (Fig. 7D). According to CTD, among the diseases we analyzed in this study, valproic acid is a therapeutic drug for anxiety, autism, bipolar disorder, and schizophrenia (Fig. 7E). This drug is also an efficient anti-convulsant used to treat epilepsy (Tomson et al., 2016) because it facilitates gamma-aminobutyric acid (GABAergic) neurotransmission (Chateauvieux et al., 2010). There is extensive evidence in the literature of the anti-inflammatory effects of valproic acid and its potential use to treat conditions such as spinal cord injury (S. Chen et al., 2018), renal ischemia (Costalonga et al., 2016), and sepsis-induced heart failure (Shi et al., 2019). Valproate was also speculated as a potential repurposing candidate to treat diseases caused by infectious agents, such as COVID-19 (Pitt et al., 2021) and toxoplasmosis (Goodwin et al., 2008). HDAC inhibitors promote epigenetic modifications in the genome that induce the expression of genes in many biological functions and cell types (Hull et al., 2016). This could explain valproic acid’s versatility and why it ranked first in our analysis.

Among the other top 20 drugs, we found molecules that are currently under investigation for repositioning from one disease category to another. Methotrexate (Fig. 7D), which affects 141 genes among the 345 hubs, is used to treat several inflammatory diseases, including psoriasis, lupus, and arthritis (Fig. 7E). Recently, a randomized clinical trial revealed a potential for methotrexate to treat positive symptoms in schizophrenia patients (Chaudhry et al., 2020). The authors of the trial argue that this effect of methotrexate might be achieved through resetting of systemic regulatory T-cell control of immune signaling, which is also the way this drug is thought to act in autoimmune diseases (Chaudhry et al., 2020). The use of anti-inflammatory drugs for the treatment of neuropsychiatric diseases gained traction in recent years (Kohler et al., 2016; Ozben and Ozben, 2019; Pandurangi and Buckley, 2020; Rosenblat et al., 2016) influenced by the increasing evidence that these disorders have underlying immune causes, which we have extensively demonstrated in this study. Dexamethasone (Fig. 7D) is a glucocorticoid anti-inflammatory drug listed in CTD as a therapy for arthritis and depression (Fig. 7E), but it is also used to treat several other inflammatory disorders. Indeed, dexamethasone was one of the few drugs submitted to randomized clinical trials that reduced mortality in COVID-19 patients subjected to invasive ventilation (RECOVERY Collaborative Group, 2021). Several of the other top 20 drugs were also listed in CTD to be used as therapy for diseases of different categories, such as cyclosporine (hepatitis C and psoriasis), indomethacin (Alzheimer’s and autoimmune diseases), dronabinol (neuropsychiatric diseases and HIV infection), and quercetin (anxiety and influenza; Fig. 7E).

## Discussion

Similar to the exponential increase in the number of published papers seen in the past decades (Fortunato et al., 2018), the number of genes associated with psychiatric, inflammatory, and infectious diseases have also increased significantly in the past 30 years . This rapid growth in knowledge about the genetic underpinnings of these diseases can be directly attributed to at least two historical landmarks: the publication of the human genome in 2001 (Lander et al., 2001; Venter et al., 2001) and the advent of high-throughput DNA-sequencing technologies (Margulies et al., 2005). Discrete advances in genes associated with specific diseases could also be spotted throughout the period analyzed here. In 1996, the triple therapy for HIV was developed using nucleoside reverse-transcriptase inhibitors and protease inhibitors (Hammer et al., 1996). In the same year, 50% of the new genes added to the knowledge network were connected to HIV infection. In 2005, a peak of novel genes associated with psoriasis and systemic lupus erythematosus was detected. This year also saw the discovery of the Th_17_ cell lineage (Langrish et al., 2005). The central role of these pro-inflammatory cells in the pathogenesis of autoimmune and infectious diseases was later identified (Zambrano-Zaragoza et al., 2014). Indeed, the key genes of the differentiation and maintenance of the Th_17_ phenotype in CD4^+^ T lymphocytes, such as interleukin 17F (IL17F), interleukin 21 (IL21), the peroxisome proliferator-activated receptor gamma (PPARG), and the fatty acid-binding protein 5 (FABP5), were connected to psoriasis and systemic lupus erythematosus in the network in 2005 (Hwang, 2010; Nalbant and Eskier, 2016).

One of the advantages of using text mining and network medicine to study the relationships between genes and diseases is the possibility of detecting novel connections from established scientific knowledge. When two diseases share a genetic mechanism, they can also present common clinical or epidemiological characteristics, despite having distinct etiological backgrounds (Barabási et al., 2011). These similarities can inform researchers of potential treatment options (Lüscher Dias et al., 2020). Here, we showed that diseases from inflammatory, psychiatric, and infectious etiologies significantly share genes with each other. This sharing was strong between disease pairs that were well studied together, such as depression and fibromyalgia. Conversely, the gene sharing between psoriasis and malaria could be perceived in our knowledge networks since the 2000s, but the number of papers featuring the two conditions together in PubMed is virtually null. We detected a few such cases, mostly involving neglected infectious diseases, which could explain the knowledge gap. We also found cases of diseases that just recently began to share genes that also lack many publications directly connecting them in the literature. A case in point is autism and RSV. We also found disease pairs, such as dementia and *Toxoplasma gondii* infection, for which there have been direct associations in the literature since 1990, but that just recently started to share genes in the network. Our results reveal potentially underexplored pathways for future research on the association between diseases of distinct categories and also for the discovery of new genes related to well-studied disease pairs.

The sharing of genes between diseases from distinct categories also reflects in the overlap of biological functions, particularly those related to immunological processes. The genes of several diseases in all categories presented enrichment for Reactome pathways related to the interferon response, cytokines, and NFkB-mediated inflammation. This pattern was detectable in our networks since the early 1990s for inflammatory diseases and gradually appeared for infectious and psychiatric diseases as well. Pathways associated with neurotransmission were almost exclusively enriched among the genes of psychiatric diseases.

Nevertheless, we found enrichment for a neurotransmission-related pathway, “transcriptional regulation by MECP2”, among the genes of HIV infection and hepatitis B that could point to a connection between these disorders and Rett syndrome, a neurological condition. Our functional enrichment results also highlighted the relevance of core cellular functions in diseases of all categories, such as signal transduction and the regulation of gene expression by transcription factors.

Our network medicine text mining approach also revealed how shared genes between disease categories can signal toward common therapeutic solutions. The findings presented in the last section of our study emphasize the relevance of drugs that target shared genes for the treatment of distinct diseases. Our results show that the genes targeted by therapeutic drugs shared by inflammatory, psychiatric, and infectious diseases have been associated with these disorders early in the past 30 years of scientific research. These genes are associated with inflammation, the cell cycle, apoptosis, and central pathways of cellular function. We also demonstrated that well-established and promising cases of repositioning involve drugs that target shared genes between diseases. Future studies should aim to reveal more common molecular mechanisms between these categories of diseases as well as to harness that knowledge for novel drug discovery and repurposing.

In summary, we could apply a machine learning and cognitive computing text-mining strategy using WDD to extract knowledge about genes related to inflammatory, infectious, and psychiatric diseases from the scientific literature and depict how this knowledge evolved during the past 30 years.

## Methods

### Knowledge network construction

We built knowledge networks containing interactions between diseases and genes using the WDD (Y. Chen et al., 2016). WDD discovers connections between genes and diseases using a natural language processing algorithm that reads full texts from PMC open access journals, patents, and abstracts in the MEDLINE (PubMed) database. A connection is found when two terms of interest (e.g., genes and diseases) are detected in the same sentence, separated by a preposition or a verb. These connections can be derived from many sources of evidence, such as gene expression, disease-associated mutations, genome-wide association studies, or protein expression experiments. WDD attributes a confidence score (0–100%) to each association based on the number of documents in which the relation is found and also on the semantic relevance of the link, determined by the natural language processing algorithm.

We performed independent searches on WDD with 27 inflammatory diseases, 63 infectious diseases, and 9 psychiatric and neurological disorders (Table S1) in July 2018. WDD returned lists of genes related to these diseases according to the scientific literature in each year from 1990 to 2018. These associations are cumulative, that is, the genes associated with the diseases in 2018 include all the associations present in the previous year. We only kept connections between genes and diseases supported by a confidence score of at least 50% and 2 documents of evidence. Custom R code was used to process, filter, and analyze data and to plot figures. The full code of all analyses and figures in this study is available at https://github.com/csbl-usp/evolution_of_knowledge.

### Evolution of knowledge

We calculated Fisher’s exact test p-value of the gene overlap between each pair of diseases in each year from 1990 to 2018. The total number of genes connected in the network in each year was used as Fisher’s exact test universe. For each year, a disease-disease knowledge network was developed using the –log_10_pval of the Fisher’s exact test (“disease-disease similarity”) as the edge weight for each disease pair. The networks were constructed using the R package *igraph* (Csardi and Nepusz, 2006) and plotted using the package *ggraph*. We detected new genes in each year by comparing the list of genes of the diseases in one year to the list of genes of the same disease in the previous year. Thus, we obtained a list of new genes that were added to the network in each year from 1991 to 2018. The total number of genes associated with each disease was also calculated for each year. Line, violin, and ridge plots were created to illustrate the results using *ggplot2* (Wickham, 2016).

### Evolution of disease relationships between categories

For the top 9 diseases of each category that were connected to the most genes in 2018 (“top 9 diseases”), we detected the diseases from the other two categories with the most significant gene sharing between them (“disease pairs”) and analyzed how these relationships evolved from 1990 to 2018. The disease-disease similarity scores obtained previously were also used in this analysis. We used the *MeSH.db* R package (Tsuyuzaki et al., 2015) to obtain the MeSH IDs and terms of all 99 diseases. Using the obtained MeSH terms of the diseases in each pair, we used the *easyPubMed* R package to search for PubMed papers in which both disease MeSHes were found together. We then used an adapted version of the *fetch_pubmed_data* function (see code in GitHub) of the *easyPubMed* package to retrieve the number of papers that contained the searched MeSH pairs in each year from 1990 to 2018. We used the disease-disease similarity score and the number of papers in 2018 to calculate a similarity-to-paper ratio for each disease pair as follows:

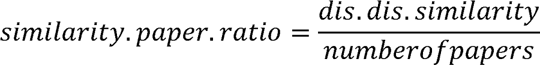

Low similarity-to-paper ratios (<10) were considered as cases of low knowledge gap between the gene sharing and the general scientific interest in the disease pairs. Pairs in this category included those in which the diseases did not share a significant amount of genes or pairs of similar diseases for which there is also a proportional number of papers that cite the two diseases together. Ratios between 10 and 40 were considered as cases of intermediate knowledge gap, that is, the diseases in the pair are similar in the genes they share, but the number of papers on the two diseases together is not proportionally high. High similarity-to-paper ratios (>40) were interpreted as cases of a large knowledge gap. The pairs that fell in this category include diseases that share a significant proportion of their genes but that have almost never been studied together, evidenced by the very low number of papers including the two MeSH terms.

### Evolution of biological pathways

We used the *enricher* function of the R package *clusterProfiler* (Yu et al., 2012) to perform an ORA against Reactome pathways of the genes associated with the top 9 diseases of each category in each year. We selected the significant Reactome pathways (p.adjust < 0.01) of the top 9 diseases in 2018 and calculated the significance of the gene overlap between these pathways with Fisher’s exact test. We considered only the genes of each significant pathway that were also present in the 2018 gene-disease network. By doing this, we limited pathways to cluster according to the genes shared from our data set, not all the genes in the pathways. We then built a pathway network connecting the significant Reactome terms using the –log_10_pvalue of the Fisher’s exact tests as edge weights, similar to what was done for the disease-disease network in Fig. 1A. We detected clusters of pathways in this network using the *cluster_louvain* function (Blondel et al., 2008) of the *igraph* R package (Csardi and Nepusz, 2006). Edge weights were considered for the cluster detection. We calculated the weighted degree of each pathway in the network using the *strength* function of the *igraph* package (Csardi and Nepusz, 2006). We manually annotated the detected clusters for their major biological function using the pathways with the highest weighted degree in each cluster as reference. The significance values (–log_10_pval) of ORA for the pathways in each cluster were used to make box and ridge plots to illustrate the results for each disease in 2018 and how these results changed from 1990 to 2018.

### Evolution of drug target hub genes

Using the 2018 gene-disease network, we detected the genes common to all three categories of diseases (“hub genes”). We used the R package *UpsetR* to visualize the number of genes shared and exclusive to the disease categories. We downloaded the drug-gene and the drug-disease interaction databases from the CTD (http://ctdbase.org/; Davis et al., 2021). We used the MeSH terms of the 99 diseases to filter the drug-disease database and kept only interactions between drugs and diseases that were listed as “therapeutic” by CTD. These are cases of a “chemical that has a known or potential therapeutic role in a disease (e.g., chemical X is used to treat leukemia)”, according to the CTD glossary (Davis et al., 2021). We filtered the drug-gene database and kept only the interactions between the therapeutic drugs and the hub genes of our analysis. This final drug-gene list was used to detect the top 20 drugs that target the most hub genes and the top 20 hub genes most targeted by the therapeutic drugs. We visualized these drug-gene interactions in a network built with the R packages *igraph* and plotted with *ggplot2* and *ggraph*. We used the yearly gene-disease networks to detect when the top 20 drug target hub genes were first connected to diseases in each category to build a timeline.

### Competing interests

We declare that the authors have no conflicts of interest.

### Data availability

The data and code used to produce the analyses and figures in this study are available at https://github.com/csbl-usp/evolution_of_knowledge.

## Author Contribution

Conceptualization, Investigation, Data Curation and Writing: TLD, RJSD, PPA, VS, GRF and HIN. Software Programming, Formal analysis: TLD, VS, TLA. Repository was developed by TLD and TLA. Resources, Writing Review & Editing: TLD, RJSD, PPA, VS, GRF and HIN; Supervision and Funding acquisition: GRF and HIN.

## Funding

This work was supported by Brazilian National Council for Scientific and Technological Development (grant numbers 313662/2017-7); the São Paulo Research Foundation (grant numbers 2018/14933-2).

---

**Figure S1.**
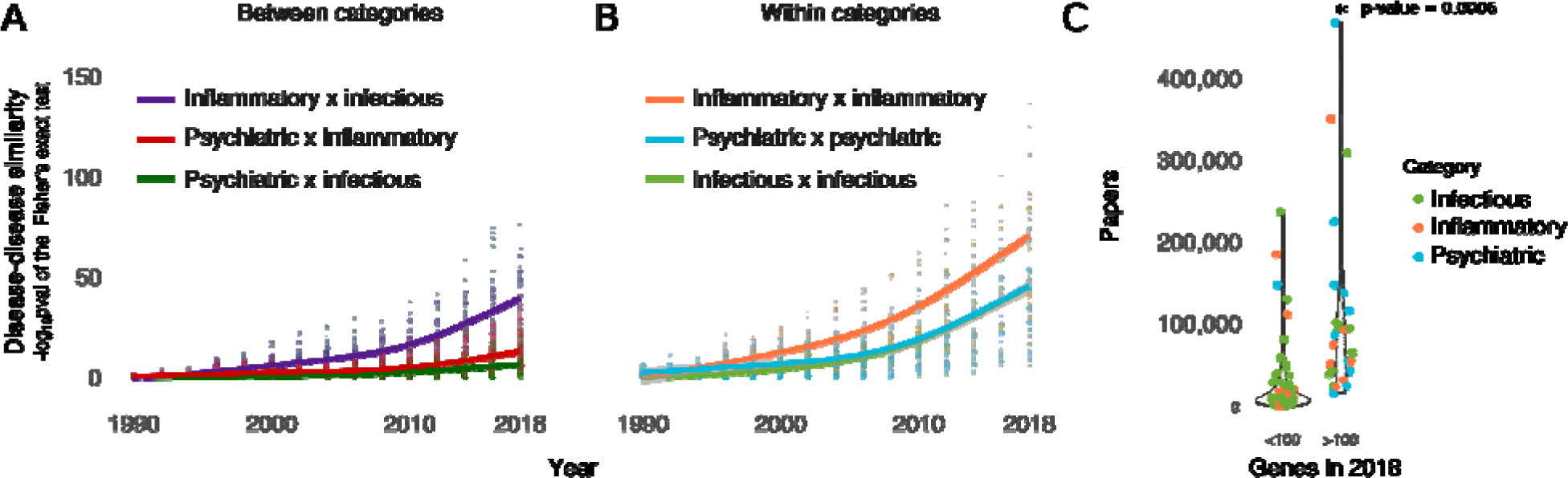
Evolution of knowledge – supplementary results. **A and B.** Evolution of the mean disease-disease similarity between diseases of different categories (**A**) or within diseases of the same categories (**B**). **C.** Comparison of the total number of papers retrieved from PubMed on diseases of all categories that were connected to less than 100 genes in the 2018 network with that connected to more than 100 genes in 2018. The *p*-value is obtained from the t-test of the mean comparison between the two distributions.

**Figure S2.**
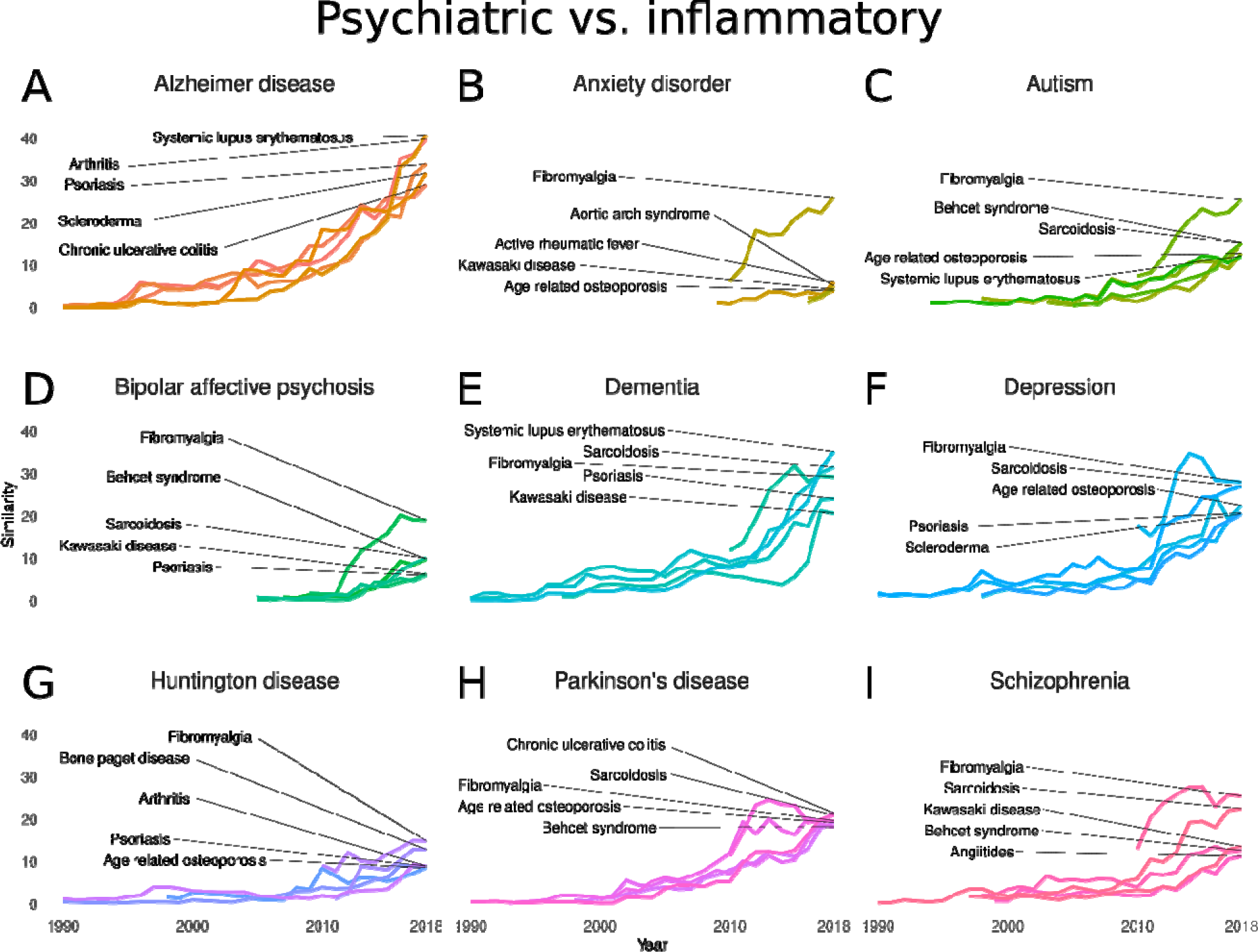
Disease-disease similarity evolution between psychiatric and inflammatory diseases from 1990 to 2018. **A–I.** Evolution of the similarity between psychiatric disorders and inflammatory diseases: Alzheimer’s disease (**A**), anxiety disorder (**B**), autism (**C**), bipolar disorder (**D**), dementia (**E**), depression (**F**), Huntington’s disease (**G**), Parkinson’s disease (**H**), and schizophrenia (**I**). Similarity scores represent the –log_10_pval of the Fisher’s exact test result of the gene overlap between each disease pair in each year from 1990 to 2018.

**Figure S3.**
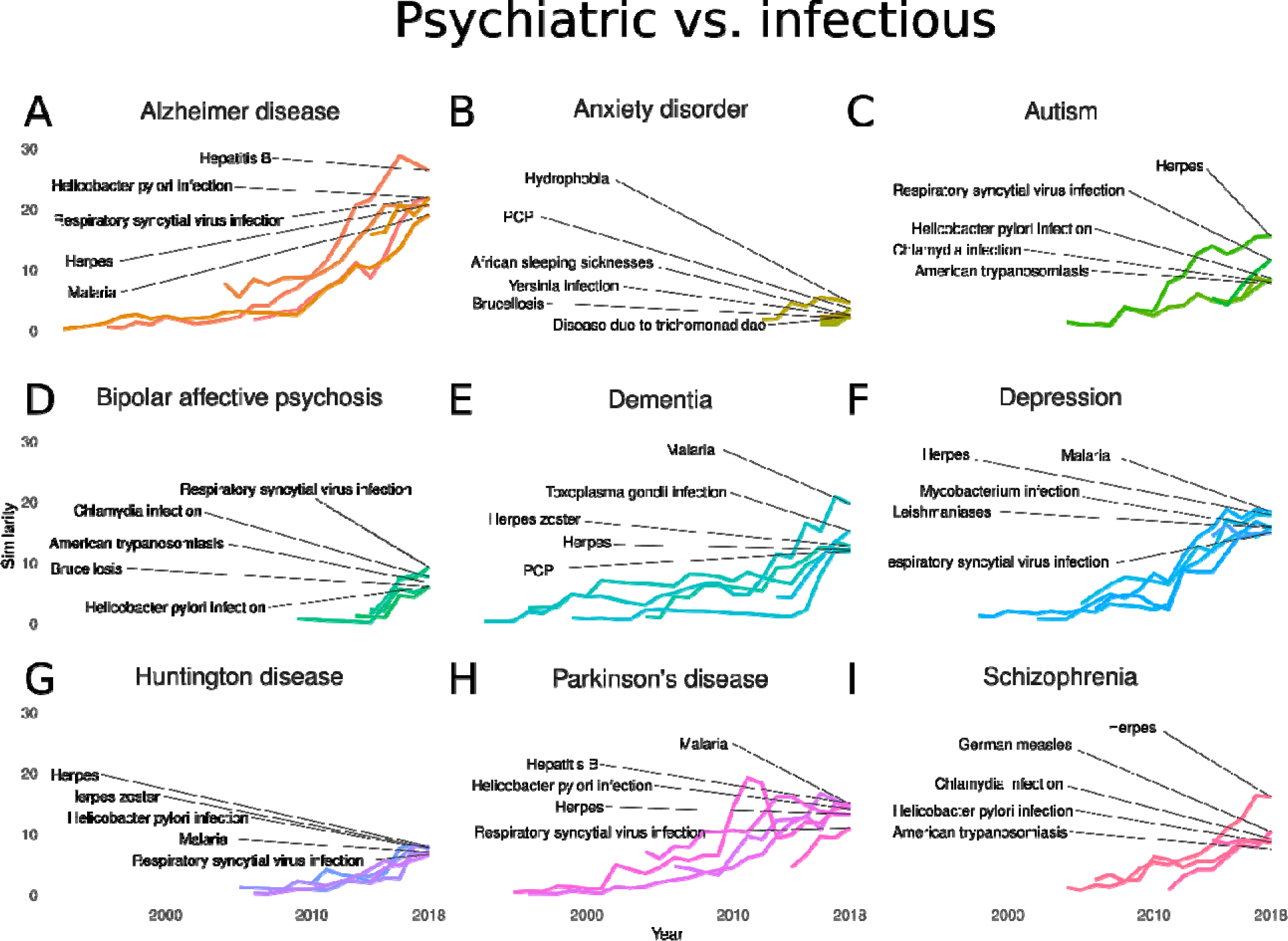
Disease-disease similarity evolution between psychiatric and infectious diseases from 1990 to 2018. **A–I.** Evolution of the similarity between psychiatric disorders and infectious diseases: Alzheimer’s disease (**A**), anxiety disorder (**B**), autism (**C**), bipolar disorder (**D**), dementia (**E**), depression (**F**), Huntington’s disease (**G**), Parkinson’s disease (**H**), and schizophrenia (**I**). Similarity scores represent the –log_10_pval of the Fisher’s exact test result of the gene overlap between each disease pair in each year from 1990 to 2018.

**Figure S4.**
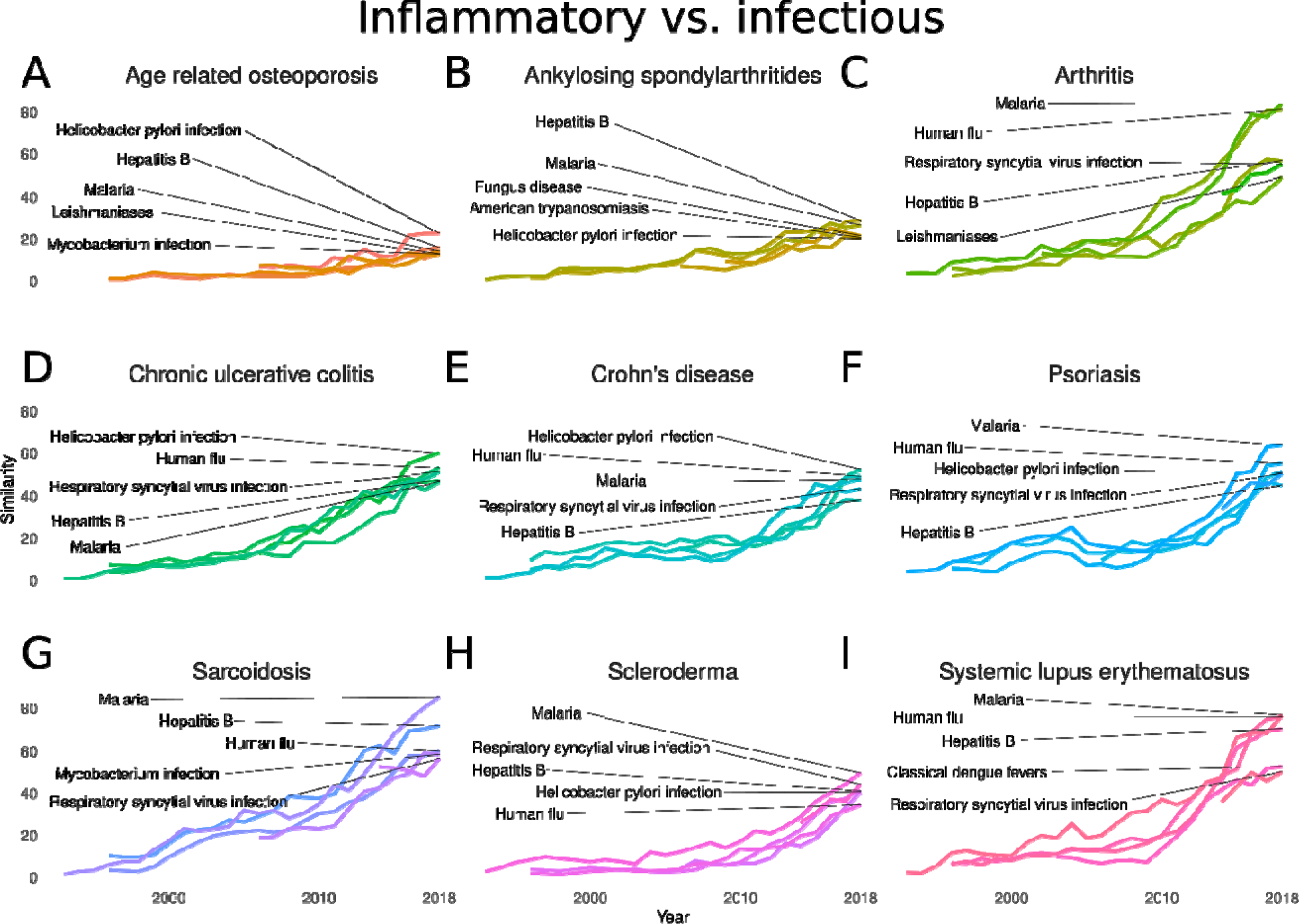
Disease-disease similarity evolution between inflammatory and infectious diseases from 1990 to 2018. **A–I.** Evolution of the similarity between psychiatric disorders and infectious diseases: age related osteoporosis (**A**), ankylosing spondylarthritides (**B**), arthritis (**C**), chronic ulcerative colitis (**D**), Crohn’s disease (**E**), psoriasis (**F**), sarcoidosis (**G**), scleroderma (**H**), and systemic lupus erythematosus (**I**). Similarity scores represent the –log_10_pval of the Fisher’s exact test result of the gene overlap between each disease pair in each year from 1990 to 2018.

**Figure S5.**
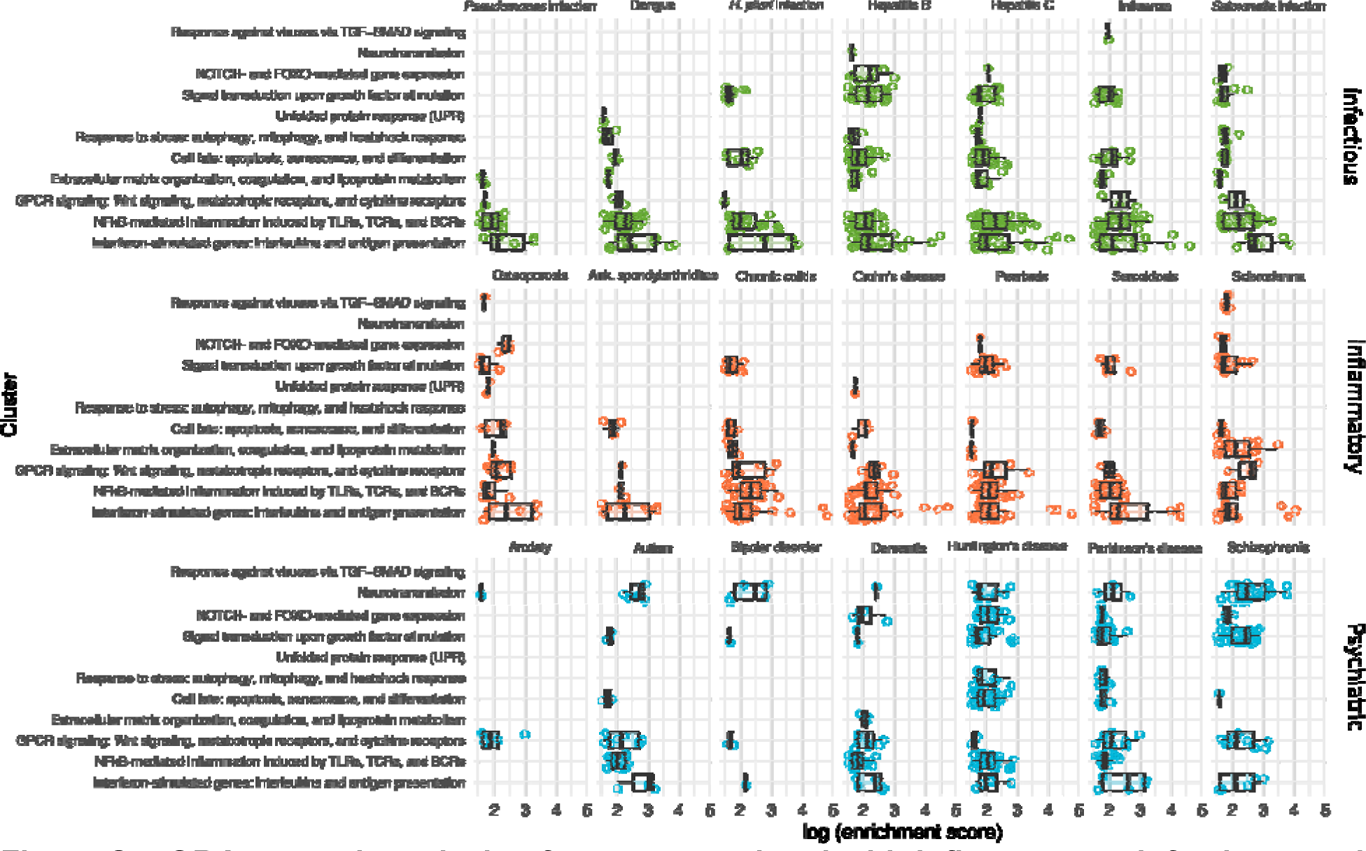
ORA network analysis of genes associated with inflammatory, infectious, and psychiatric diseases. Enrichment score distribution of the terms in the clusters from Fig. 4 for diseases not depicted in Fig. 5. Box plots illustrate the distribution of the enrichment scores of the Reactome pathways in each cluster.

## Notes

### Competing Interest Statement

The authors have declared no competing interest.

### Clinical Trial

Not applicable

